# COVID-19 risk factors amongst 14,786 care home residents: An observational longitudinal analysis including daily community positive test rates of COVID-19, hospital stays, and vaccination status in Wales (UK) between 1^st^ September 2020 and 1^st^ May 2021

**DOI:** 10.1101/2021.09.30.21264338

**Authors:** Joe Hollinghurst, Robyn Hollinghurst, Laura North, Amy Mizen, Ashley Akbari, Sara Long, Ronan A Lyons, Rich Fry

**Affiliations:** Swansea University; Open University, Swansea University; Population Data Science, Swansea University

## Abstract

**Objectives:** Determine individual level risk factors for care home residents testing positive for SARS-CoV-2.

**Study Design:** Longitudinal observational cohort study using individual-level linked data.

**Setting:** Care home residents in Wales (United Kingdom) between 1st September 2020 and 1st May 2021.

**Participants:** 14,786 older care home residents (aged 65+). Our dataset consisted of 2,613,341 individual-level daily observations within 697 care homes.

**Methods:** We estimated odds ratios (ORs [95% confidence interval]) using multilevel logistic regression models. Our outcome of interest was a positive SARS-CoV-2 polymerase chain reaction (PCR) test. We included time dependent covariates for the estimated community positive test rate of COVID-19, hospital admissions, and vaccination status. Additional covariates were included for age, positive PCR tests prior to the study, sex, frailty (using the hospital frailty risk score), and specialist care home services.

**Results:** The multivariable logistic regression model indicated an increase in age (OR 1.01 [1.00,1.01] per year of age), community positive test rate (OR 1.13 [1.12,1.13] per percent increase in positive test rate), hospital inpatients (OR 7.40 [6.54,8.36]), and residents in care homes with non-specialist dementia care (OR 1.42 [1.01,1.99]) had an increased odds of a positive test. Having a positive test prior to the observation period (OR 0.58 [0.49,0.68]) and either one or two doses of a vaccine (0.21 [0.17,0.25] and 0.05 [0.02,0.09] respectively) were associated with a decreased odds of a positive test.

**Conclusions:** Our findings suggest care providers need to stay vigilant despite the vaccination rollout, and extra precautions should be taken when caring for the most vulnerable. Furthermore, minimising potential COVID-19 infection for care home residents admitted to hospital should be prioritised.

**SUMMARY BOXES:** *Section 1: What is already known on this topic:*

- Care home residents are at a high risk of COVID-19 infection, but existing literature has mainly focussed on excess mortality rather than infection risk.
- In our study we were able to investigate associations between COVID-19 infections and the community positive test rate of COVID-19, the vaccination status of care home residents, hospital admissions, and frailty.

*Section 2: What this study adds:*

- Our study suggests an increased community positive test rate and hospital inpatients had an increased likelihood of a positive SARS-CoV-2 polymerase chain reaction test, whilst one or two doses of vaccination indicated a decreased chance of a positive test.
- Our findings suggest care providers need to stay vigilant despite the vaccination rollout, and extra precautions should be taken when caring for the most vulnerable, especially in a hospital setting.

## INTRODUCTION

Care homes are a keystone of adult social care. They provide accommodation and care for those needing substantial help with personal care, but more than that, they are people’s homes (1,2). In 2016, there were 11,300 care homes in the UK, with a total of 410,000 residents (3). Within care homes people live in proximity, and may live with frailty and many different health conditions, making them susceptible to outbreaks of infectious disease (1). COVID-19 is described by Lithande et al, as ‘…a dynamic, specific and real threat to the health and well-being of older people’ (2020,p.10) (4). The impacts of COVID-19 on this sub-population have been reported widely in both international and UK media, and in a growing peer reviewed literature.

Vaccinations for SARS-CoV-2 in the UK have been prioritised for those identified at higher risk including to older people living in care homes (5,6). However, very few vaccination trials have recruited older people or older people with frailty (7). Most studies involving care homes have used data sources at an aggregated residential-level, rather than individual-level. This has included the risk of outbreaks of COVID-19 following hospital discharges (8), along with increased risk of SARS-CoV-2 infection due to differing levels of community prevalence (9). Individual-level analyses have focussed on mortality due to COVID-19 in care homes, rather than the risk factors for individuals being infected (10–12).

This is the first study we are aware of investigating this vulnerable sub-population at an individual-level with the inclusion of the community positive test rate of COVID-19, hospital admissions, and vaccination status. Furthermore, we included information on previous positive SARS-CoV-2 polymerase chain reaction (PCR) tests, age, sex, and frailty. As suggested in (13) we were able to do this using up-to-date linked data on care homes from the Secure Anonymised Information Linkage (SAIL) Databank (14–16).

## Objectives

We aimed to identify individual level risk factors for SARS-CoV-2 infection with the inclusion of community positive test rate of COVID-19, hospital admissions, and vaccination status.

## METHODS

### Study design

Longitudinal observational cohort study using anonymised linked data from the Secure Anonymised Information Linkage (SAIL) Databank.

### Participants and Setting

Our cohort was 14,786 older care home residents (aged 65+) living in Wales between 1^st^ September 2020 and 1^st^ May 2021. Our dataset consisted of 2,613,341 individual level daily observations for the same period within 697 care homes. Residents were included if they lived in a care home at any period during the observation window. Only residents with at least one PCR test (positive or negative) were included within the dataset. Residents were censored if they moved out of the care home or died. All data were collected retrospectively and linked anonymously within the SAIL Databank.

### Data sources

We used linked longitudinal data from the SAIL Databank to create our dataset (14–16). We used the Welsh Demographic Service Dataset (WDSD) to determine care home residents and residency dates by linking an anonymised residential linkage field to an anonymised care home registry derived from Care Inspectorate Wales (CIW). The WDSD also contains the Lower-layer Super Output Area (LSOA) for each address, which is an area containing approximately 1,500 people, as well as demographic information (age and sex). We used the COVID Vaccine Dataset (CVVD) to identify when individuals had received their vaccinations. The Pathology COVID-19 Daily (PATD) data was used to identify dates of positive and negative SARS-CoV-2 PCR tests. We linked to the Patient Episode Database for Wales (PEDW) to include an indicator for hospital admissions and to calculate the hospital frailty risk score. We used a combination of the Office for National Statistics (ONS) Annual District Death Extract (ADDE), WDSD, and Consolidated Death Data Source (CDDS) for mortality information for censoring.

### Variables

#### Outcome - positive SARS-CoV-2 PCR test

Our outcome of interest was a positive SARS-CoV-2 PCR test. The date of the positive test was recorded as the date when the specimen was collected. We used a binary variable to indicate the positive test dates for individuals.

#### Exposures

Exposure variables were daily COVID-19 community positive test rate estimates and a daily indicator for if a resident was a hospital inpatient. Daily inpatient status was identified using PEDW, residents were identified as being a hospital inpatient for all dates from the admission to the discharge date (inclusive) of a hospital spell.

The estimated COVID-19 community positive test rate was calculated by removing all tests for care home residents and creating a geospatial model for each daily observation. The model includes a spatial correlation term that decays with distance. In other words, areas that are close in proximity are likely to have a more similar estimate than those further away. This reduces the impact of artificial boundaries introduced by statistical geographies and provides a more realistic spatial distribution estimate of positive test rates in the communities surrounding a care home. The community test rate estimates were calculated for each LSOA using a 14-day lookback window for each date. The testing strategy in Wales in the observation period included asymptomatic testing of those caring for vulnerable people (e.g. care home workers and healthcare workers), and symptomatic testing for those in the community (17). Our community positive test rate estimate included both symptomatic and asymptomatic testing. The geospatial model used is an extension of the logistic regression model for binomial (numerator/denominator) data, in which the log-odds of the probability, *P(x)*, of at least one positive PCR test is the unobserved realisation of a spatially correlated stochastic process. Specifically, we used the total number of positive PCR tests in the 14-day window as the numerator, and total tests in the 14-day window as the denominator. The model has three parameters that determine the mean and variance of *P(x)* and the rate at which the correlation between the values of *P(x)* at two different locations decays with increasing distance between them, for more detail on the methodology see (18).

#### Predictors and Confounders

We included the number of doses of a vaccine and a binary indicator for if an individual had a positive PCR test prior to the observation period as predictor variables. The number of vaccine doses received was time varying and was recorded as a categorical variable on each date (0 doses, 1 dose, 2 doses). Additional variables included were specialist services provided by the care home where individuals were resident during the study period. This included: nursing care (yes/no), learning disability (yes/no), mental health (yes/no), dementia (no or unknown, non-specialist, specialist). Age (continuous), sex (male/female) and hospital frailty risk score (no score, low, intermediate, high) were included at the individual level. The Hospital Frailty Risk Score (HFRS) was developed using Hospital Episode Statistics (HES), a database containing details of all admissions, Emergency Department attendances and outpatient appointments at NHS hospitals in England, and validated on over 1-million older people using hospitals in 2014/15 (19). The HFRS uses the International Classification of Disease version 10 (20) (ICD-10) codes to search for specific conditions from secondary care. A weight is then applied to the conditions and a cumulative sum is used to determine a frailty status of: Low, Intermediate or High. We additionally included a HFRS score of ‘No score’ for people who had not been admitted to hospital in the look back period. We calculated the HFRS using PEDW, the Welsh counterpart to HES, on each daily observation, with a 2-year look back of all hospital admissions recorded in Wales on each date.

### Longitudinal dataset design

In the longitudinal dataset created for the analysis individuals have multiple daily observations. Each daily observation includes updated time-dependent covariates; the estimated community COVID-19 positive test rate, whether an individual was in hospital, and the number of vaccine doses received. All other variables were fixed at the value of entry into the study. Additionally, the dataset has anonymous individual and care home identifiers used to cluster the observations. For an example of the dataset design see Figure S1 and Table S1.

### Bias

To help minimise selection bias we only included individuals who had at least one SARS-CoV-2 PCR test. Similarly, to minimise time interval bias we used a large observation period where we believe testing of care home residents was more consistent than at the start of the pandemic. We also included anonymous individual and care home level identifiers to account for correlation amongst repeated measures.

### Statistical methods

Descriptive statistics included the demographic information for individuals at the start of their residency and stratifications for those who did and did not have a positive PCR test within the observation period. We produced time plots for the daily estimated community positive test rate, positive test rate of residents, numbers of PCR tests for residents, and number of positive PCR tests for residents. For our analysis we used multilevel logistic regression models with a random intercept term for each care home. The regression was applied to the individual-level daily dataset. For sensitivity analysis we calculated null (intercept only) models with random effects at the individual level, care home level, and both (see Table S1). Observations with a missing LSOA were removed, and individuals were censored if they moved residence or died.

### Patient and public involvement

No patients or members of the public were involved in setting the research question, study design, outcome measures, or the conduct of the study.

## RESULTS

### Participants and descriptive data

We analysed 2,613,341 daily observations, consisting of 14,786 care home residents within 697 care homes. The reasons for exclusion from the analysis dataset are included in Figure 1. Descriptive statistics taken from the first observation for each individual are included in Table 1. We observed a high mean age of 85 (standard deviation 8.2), a large proportion of females (69%), and a high proportion of individuals with frailty (61.8% with low, intermediate, or high HFRS). To provide an indication of demographic differences we also stratified the variables by those who did and did not have a positive SARS-CoV-2 PCR test at any point within the observation period.

**Figure 1.**
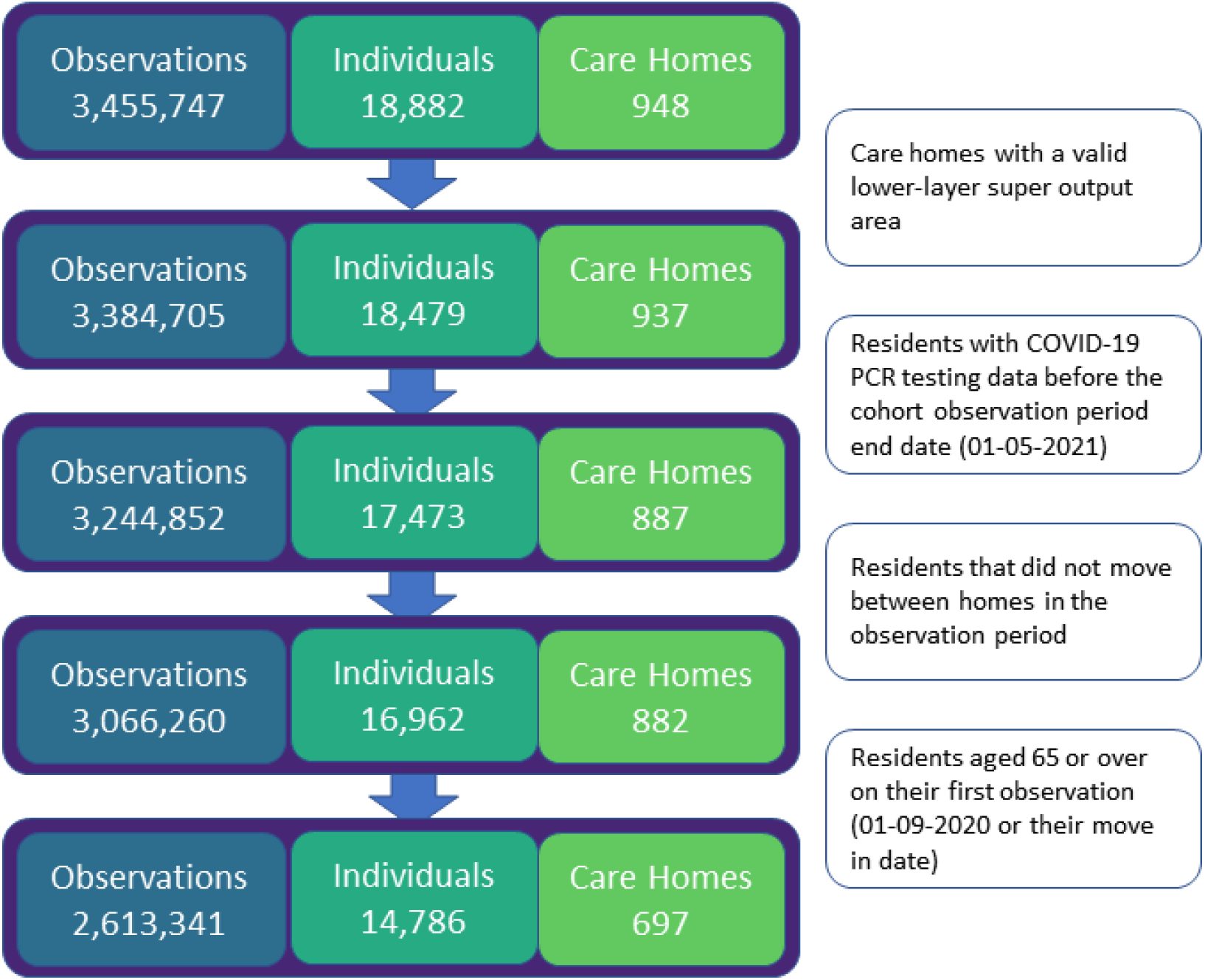
Flow diagram for the number of observations, individuals, and care homes in the study.

**Table 1.** Descriptive statistics for care home residents at the start of the observation period (1^st^ September 2020, or first recorded observation).

|  | All |  | Negative |  | Positive |  |
| --- | --- | --- | --- | --- | --- | --- |
| **Individuals (N)** | 14,786 |  | 12,629 |  | 2,157 |  |
| **Individuals with a positive test during the observation period** | 2,157 | 14.6% |  |  |  |  |
| **Care homes (N)** | 697 |  | 684 |  | 289 |  |
| **Mean age (S.D)** | 85 (8.2) |  | 85 (8.2) |  | 84.9 (8) |  |
| **Gender** |  |  |  |  |  |  |
| **Female** | 10,190 | 68.9% | 8,720 | 69.0% | 1,470 | 68.2% |
| **Male** | 4,596 | 31.1% | 3,909 | 31.0% | 687 | 31.8% |
| **Hospital Frailty Risk Score** |  |  |  |  |  |  |
| **No score** | 5,651 | 38.2% | 4,756 | 37.7% | 895 | 41.5% |
| **Low** | 1,209 | 30.4% | 1,029 | 30.9% | 180 | 27.6% |
| **Intermediate** | 3,430 | 23.2% | 2,944 | 23.3% | 486 | 22.5% |
| **High** | 4,496 | 8.2% | 3,900 | 8.1% | 596 | 8.3% |
| **Specialist services** |  |  |  |  |  |  |
| **Nursing - No** | 7,132 | 48.2% | 6,057 | 48.0% | 1,075 | 49.8% |
| **Nursing - Yes** | 7,654 | 51.8% | 6,572 | 52.0% | 1,082 | 50.2% |
| **Learning disabilities - No** | 12,495 | 84.5% | 10,760 | 85.2% | 1,735 | 80.4% |
| **Learning disabilities - Yes** | 2,291 | 15.5% | 1,869 | 14.8% | 422 | 19.6% |
| **Mental Health - No** | 8,275 | 56.0% | 7,307 | 57.9% | 968 | 44.9% |
| **Mental Health - Yes** | 6,511 | 44.0% | 5,322 | 42.1% | 1,189 | 55.1% |
| **Dementia care - No or Unknown** | 2,790 | 18.9% | 2,469 | 19.6% | 321 | 14.9% |
| **Dementia care - Non-specialist** | 8,708 | 58.9% | 7,349 | 58.2% | 1,359 | 63.0% |
| **Dementia care - Specialist** | 3,288 | 22.2% | 2,811 | 22.3% | 477 | 22.1% |

Cross tabulations for the observations with and without positive tests with hospital inpatients and the number of vaccine doses is displayed in Table 2. Results indicated a high proportion of observations with a positive PCR test had not been vaccinated (96%), and of those with a positive test who were unvaccinated a significant proportion were hospital inpatients (19%). A significantly larger proportion of observations with a positive PCR test were identified for hospital inpatients (0.93%) compared to not (0.08%).

**Table 2.** Time varying cross tabulation for daily observations dependent on hospital status (inpatient) and number of vaccine doses received (0,1, or 2). The number of positive tests exceeds the total number of individuals testing positive as individuals could test positive on more than one date.

|  |  | Observations with a Positive PCR test |  |  |  |
| --- | --- | --- | --- | --- | --- |
|  |  | Yes | N = 2,618 | No | N = 2,610,723 |
| **Hospital inpatient** | **Vaccine doses** | N | % | N | % |
| No | 0 | 2,018 | 77.08% | 1,544,048 | 59.14% |
| No | 1 | 74 | 2.83% | 581,521 | 22.27% |
| No | 2 | ≤6 | <0.23% | 428,945 | 16.43% |
| Yes | 0 | 495 | 18.91% | 39,410 | 1.51% |
| Yes | 1 | 24 | 0.92% | 11,665 | 0.45% |
| Yes | 2 | ≤6 | <0.23% | 5,134 | 0.20% |
| **Total** |  | 2,618 | 100% | 2,610,723 | 100% |

Figure 2 shows the daily estimated community positive test rate of COVID-19 along with the positive test rate, number of tests taken, and number of positive tests for care home residents. The estimated community positive test rate of COVID-19 was largely correlated with the positive test rate amongst care home residents, with peaks in November and January. There was a large decrease in testing and positive tests amongst care home residents after February.

**Figure 2.**
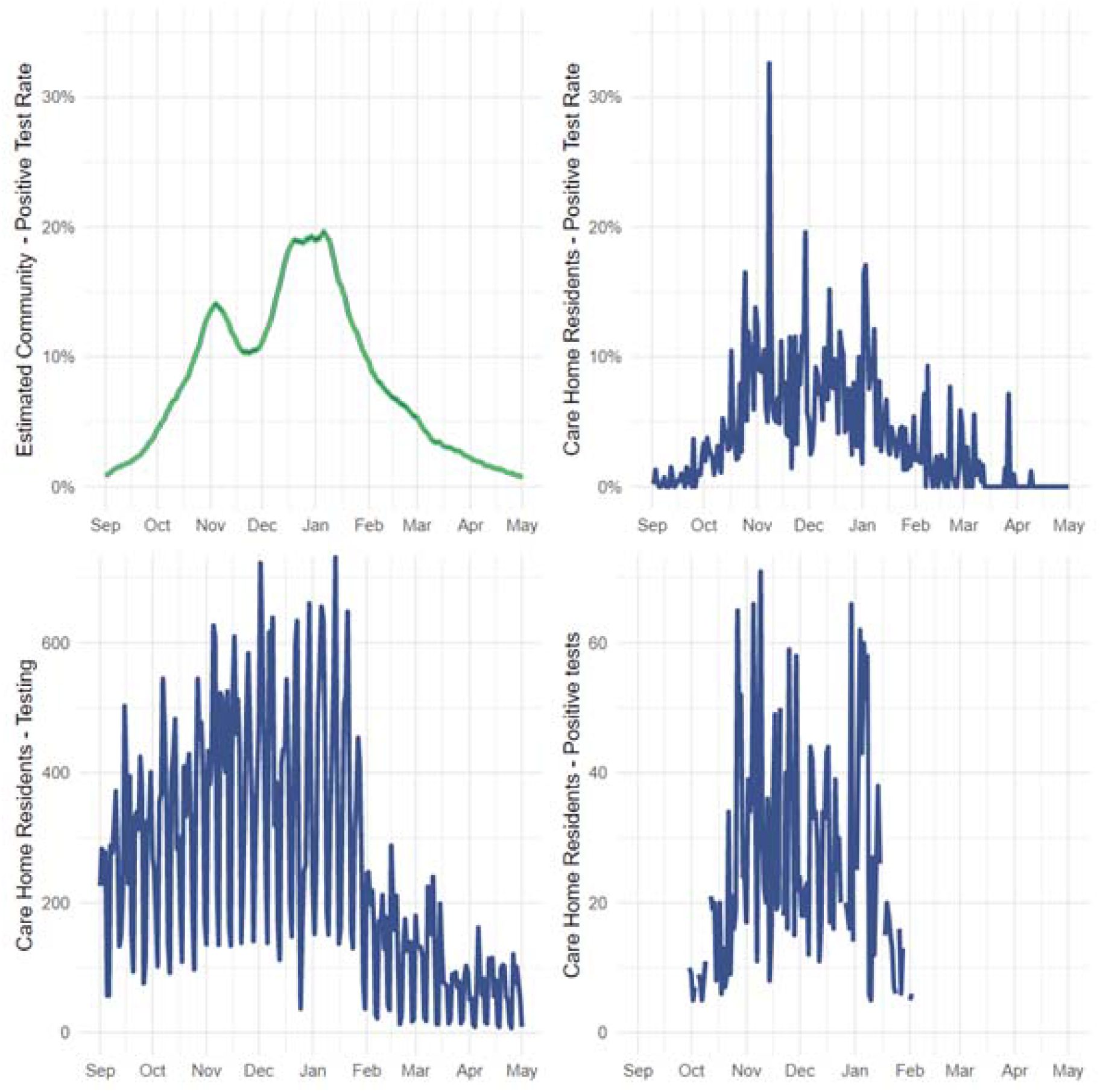
Daily estimated community positive test rate of COVID-19 (top left), positive test rate of care home residents (top right), number of tests taken for care home residents (bottom left), and the number of positive tests for care home residents (bottom right). For privacy protection the daily total of care home residents with positive tests has been masked (removed) where there were less than 5 positive tests on a particular date.

### Logistic Regression for Positive SARS-CoV-2 PCR tests

The univariable and multivariable multilevel logistic regression results for positive SARS-CoV-2 PCR tests are presented in Table 3. The multivariable model indicated an increase in age (OR 1.01 [1.00,1.01] per year of age), community positive test rate (OR 1.13 [1.12,1.13] per percentage increase in the estimated community positive test rate of COVID-19), hospital inpatients (OR 7.40 [6.54,8.36] compared to residents not in hospital), and residents in care homes with non-specialist dementia care (OR 1.42 [1.01,1.99] compared to those without specialist dementia care) had an increased odds of a positive test. Having a positive test prior to the observation period (OR 0.58 [0.49,0.68]) and either one or two doses of a vaccine (0.21 [0.17,0.25] and 0.05 [0.02,0.09] respectively compared to no vaccine) were associated with a decreased odds of a positive test. The univariable model indicated an increased odds of a positive test was also associated with sex (male compared to female OR 1.20 [1.10,1.31]), care homes with specialist mental health services (1.41 [1.06,1.86]), and increased frailty severity (ORs of 1.27 [1.09,1.47], 1.25 [1.13,1.39], and 1.48 [1.34,1.63] for low, intermediate and high hospital frailty risk scores respectively compared to no score).

**Table 3.** Univariable and multivariable multilevel logistic regression results for positive SARS-CoV-2 PCR testing amongst care home residents. Results are displayed as Odds Ratios with 95% confidence intervals.

| **Odds Ratios (95% CI)** | **Univariable** | **Multivariable** |
| --- | --- | --- |
| **Fixed effects** |  |  |
| **Age (continuous)** | **1.005 (1.000,1.010)** | **1.010 (1.004,1.015)** |
| **Sex (baseline: female)** |  |  |
| Male | 1.197 (1.099,1.305) | 1.082 (0.987,1.187) |
| **Predicted COVID-19 positive test rate % (continuous)** | **1.160 (1.153,1.167)** | **1.125 (1.118,1.133)** |
| **Positive PCR test before September (baseline: no)** |  |  |
| Positive test - yes | 0.651 (0.556,0.762) | 0.579 (0.490,0.682) |
| **Hospital inpatient (baseline: no)** |  |  |
| Hospital inpatient - yes | 9.333 (8.357,10.422) | 7.398 (6.546,8.362) |
| **Vaccine doses (baseline: no dose)** |  |  |
| 1 dose | 0.117 (0.096,0.142) | 0.206 (0.170,0.251) |
| 2 doses | 0.012 (0.006,0.024) | 0.045 (0.023,0.090) |
| **Specialist services (baseline for each category: no)** |  |  |
| Nursing care - yes | 1.084 (0.814,1.444) | 1.182 (0.885,1.579) |
| Learning disability - yes | 1.218 (0.881,1.683) | 1.109 (0.767,1.603) |
| Mental health - yes | 1.406 (1.062,1.861) | 1.252 (0.924,1.694) |
| Dementia - non specialist care | 1.469 (1.064,2.028) | 1.418 (1.014,1.985) |
| Dementia - specialist care | 1.295 (0.859,1.952) | 1.132 (0.739,1.732) |
| **Hospital Frailty Risk Score (baseline: no score)** |  |  |
| Low | 1.267 (1.090,1.472) | 1.131 (0.969,1.321) |
| Intermediate | 1.254 (1.128,1.394) | 1.054 (0.943,1.179) |
| High | 1.481 (1.343,1.633) | 1.000 (0.896,1.115) |
|  | *Null model* |  |
| **Intercept** | 0.001 (0.001,0.001) | 0 (0,0) |
| **Random effects** |  |  |
| **Variance - intercept** | 2.670 | 2.506 |
| Standard error | 0.177 | 0.176 |

The random effects term indicated significant variance at the care home level in the multivariable model (intercept variance of 2.51 with standard error 0.178). We compared intercept only (null) models with random effects terms at the individual, care home, and individual and care home level and found most of the variability was accounted for at the care home level, see Table S2.

## DISCUSSION

An increase in the community positive test rate of COVID-19 led to an increase in the odds of care home residents testing positive, with an OR of 1.13 (1.12,1.13) per percentage increase of community positive test rate. As the community positive test rate estimates ranged between 0-20% in the observed time period this suggests a potential 10-fold increase in the odds of a positive test in care home residents at the peak of community positive test rate. The association between community positive test rate and care home residents having a COVID-19 infection warrants further research with routes of potential ingress needing further exploration to provide evidence and develop robust policies. Overall this finding highlights the importance of promoting strategies that maintain lower levels of community positive test rate which help reduce the odds of infection amongst vulnerable populations.

We found care home residents who were in hospital had a large increased odds of testing positive for SARS-CoV-2 with an adjusted OR of 7.40 (6.54,8.36). This could be due to the type of hospital care home residents are admitted to, with an estimated 61.9% of COVID-19 infection being hospital acquired in residential community care hospitals, compared to 9.7% in hospitals provided acute and general care (21). Residents entering a hospital environment may be more likely to be exposed to COVID-19 due to an increased contact with healthcare workers and close proximity to other patients in wards (22). We also postulate that the increased odds of testing positive could be associated with an increased probability of being PCR tested whilst in hospital following a lateral flow test in a care home.

Increased age, sex (male), and increased frailty severity were associated with an increased odds of a positive test in the univariable models. These factors are individual level characteristics that would possibly require an increased level of care, and therefore increased daily personal contact, potentially increasing the chance of transmission of COVID-19. In the multivariable model sex and frailty were statistically insignificant, suggesting a reduced importance compared to community positive test rate and hospital stays.

Those testing positive for SARS-CoV-2 prior to the observation period and individuals with one and two doses of a vaccine were at a reduced odds of a positive PCR test, with adjusted odds ratios of 0.58 (0.49,0.68), 0.21 (0.17,0.25) and 0.05 (0.02,0.09) respectively. Those who had previously tested positive may have built up immunity to COVID-19 in a similar way to receiving a vaccine (23). The cross tabulated results and interaction models indicated the odds of a positive test were elevated for those in hospital with a vaccine compared to those not in hospital. This suggests although the vaccine may be effective, there is still an increased risk of a positive test for hospital inpatients.

### Strengths

We were able to create a large longitudinal dataset of care home residents and include linked individual-level data on hospital admissions, COVID-19 community positive test rate, vaccinations, and demographic information. This was all possible using existing data from the SAIL Databank.

### Limitations

We observed a change in the testing regime from February, where the total number of tests was reduced. We did not explicitly investigate the impact of the change in testing amongst care home residents but found the care home positive test rate remained consistent in shape and magnitude compared with the estimated community positive test rate. We were unable to link all care homes in Wales in the SAIL databank (91%, 948 of 1048). We were unable to include details on care home staff who may have tested positive for SARS-CoV-2 as data on care home staff is currently cannot be linked to specific care homes.

### Conclusion

Our findings indicate that measures taken to prevent COVID-19 from spreading to the most vulnerable in society are not completely effective. Whilst vaccination profoundly decreased the odds of testing positive for SARS-CoV-2, there was still an increased risk of infection for vaccinated individuals admitted to hospital. We also found that an increased community positive test rate of COVID-19 was associated with an increased odds of infection in care home residents. This may reflect higher risk from visitors or care staff living locally. Achieving and maintaining very high rates of vaccination in residents, health and care staff, and visitors is essential. We suggest that care providers need to stay vigilant despite the vaccination rollout, and extra precautions should be taken when caring for the most vulnerable. Follow up research is needed to evaluate the impact of vaccine waning and changing prevalence of viral strains.

## Supporting information

STROBE checklist

## Data Availability

The data used in this study are available in the SAIL Databank at Swansea University, Swansea, UK. All proposals to use SAIL data are subject to review by an independent Information Governance Review Panel (IGRP). Before any data can be accessed, approval must be given by the IGRP. The IGRP gives careful consideration to each project to ensure proper and appropriate use of SAIL data. When access has been approved, it is gained through a privacy-protecting safe haven and remote access system referred to as the SAIL Gateway. SAIL has established an application process to be followed by anyone who would like to access data via SAIL https://www.saildatabank.com/application-process.

## Acknowledgements

This work uses data provided by patients and collected by the NHS as part of their care and support. We would also like to acknowledge all data providers who make anonymised data available for research.

We wish to acknowledge the collaborative partnership that enabled acquisition and access to the de-identified data, which led to this output. The collaboration was led by the Swansea University Health Data Research UK team under the direction of the Welsh Government Technical Advisory Cell (TAC) and includes the following groups and organisations: the Secure Anonymised Information Linkage (SAIL) Databank, Administrative Data Research (ADR) Wales, NHS Wales Informatics Service (NWIS), Public Health Wales, NHS Shared Services and the Welsh Ambulance Service Trust (WAST). All research conducted has been completed under the permission and approval of the SAIL independent Information Governance Review Panel (IGRP) project number 0911.

We used the STROBE checklist to create this manuscript, an Explanation and Elaboration article discusses each checklist item and gives methodological background and published examples of transparent reporting. The STROBE checklist is best used in conjunction with this article (freely available on the Web sites of PLoS Medicine at http://www.plosmedicine.org/, Annals of Internal Medicine at http://www.annals.org/, and Epidemiology at http://www.epidem.com/). Information on the STROBE Initiative is available at www.strobe-statement.org.

