## Supplementary file for "COVID-19 risk factors amongst 14,786 care home residents: An observational longitudinal analysis including daily community positive test rates of COVID-19, hospital stays, and vaccination status in Wales (UK) between 1^st^ September 2020 and 1^st^ May 2021"

**Contents**

**Dataset design**

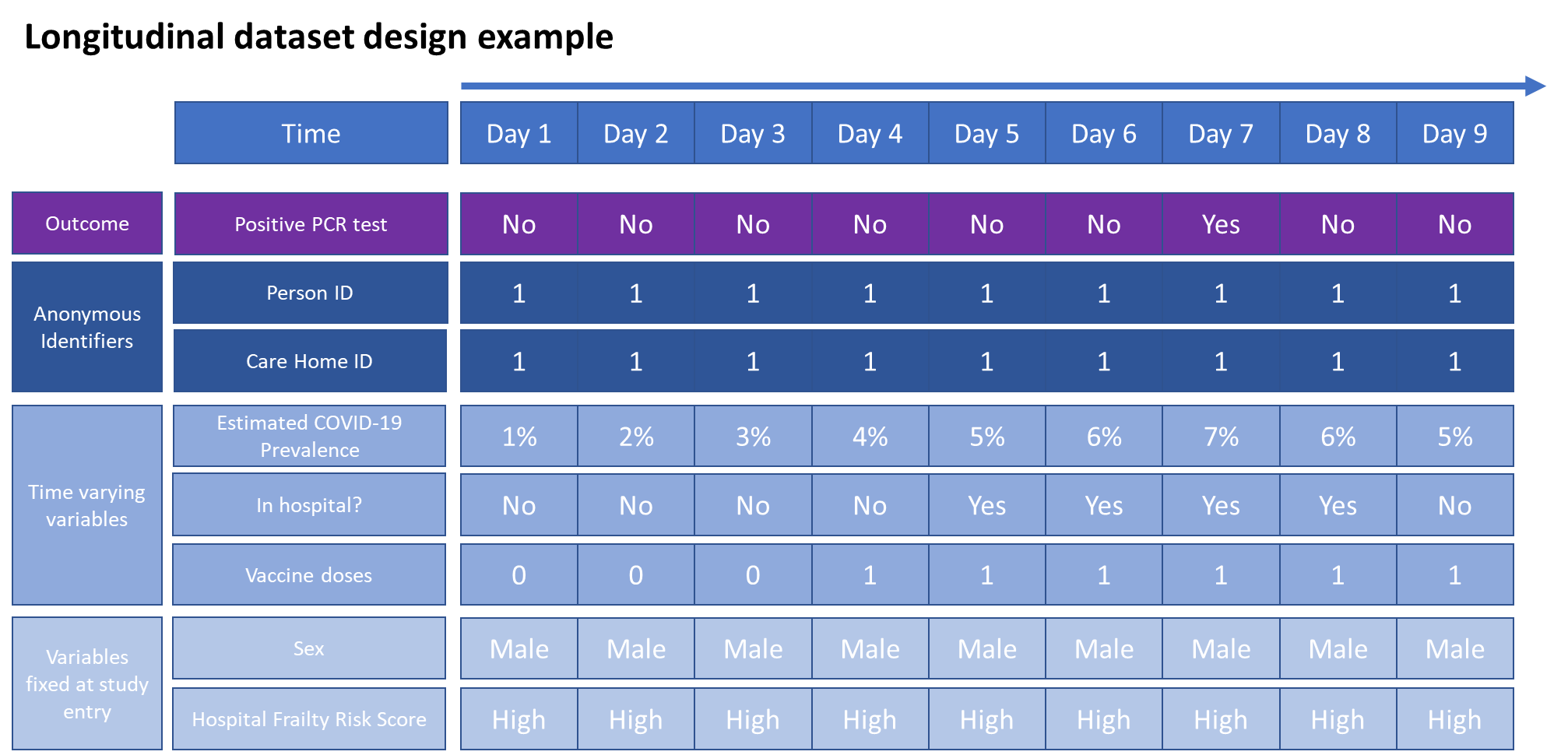

Figure S1. Fictious example of the time-dependent dataset design

Table S1. Fictitious example of the dataset used for statistical analysis.

| Person ID | Care home ID | Time | Positive PCR test | Community prevalence | Hospital Inpatient | Vaccine doses | Positive test before study start | Age | Sex | HFRS | Nursing | Learning disabilities | Mental Health | Dementia Care |
| --- | --- | --- | --- | --- | --- | --- | --- | --- | --- | --- | --- | --- | --- | --- |
| 1 | 10 | 1 | 0 | 1 | No | 0 | 0 | 70 | Male | Low | No | Yes | No | No |
| 1 | 10 | 2 | 0 | 2 | No | 0 | 0 | 70 | Male | Low | No | Yes | No | No |
| 1 | 10 | 3 | 0 | 3 | No | 0 | 0 | 70 | Male | Low | No | Yes | No | No |
| 1 | 10 | 4 | 0 | 4 | Yes | 0 | 0 | 70 | Male | Low | No | Yes | No | No |
| 1 | 10 | 5 | 1 | 5 | Yes | 0 | 0 | 70 | Male | Low | No | Yes | No | No |
| 1 | 10 | 6 | 0 | 4 | Yes | 1 | 0 | 70 | Male | Low | No | Yes | No | No |
|  |  | … |  |  |  |  |  |  |  |  |  |  |  |  |
| 2 | 11 | 1 | 0 | 1 | No | 0 | 1 | 85 | Female | High | Yes | No | No | Yes |
| 2 | 11 | 2 | 0 | 2 | No | 0 | 1 | 85 | Female | High | Yes | No | No | Yes |
| 2 | 11 | 3 | 0 | 3 | No | 0 | 1 | 85 | Female | High | Yes | No | No | Yes |
| 2 | 11 | 4 | 0 | 4 | No | 0 | 1 | 85 | Female | High | Yes | No | No | Yes |
| 2 | 11 | 5 | 0 | 5 | No | 0 | 1 | 85 | Female | High | Yes | No | No | Yes |
|  |  | … |  |  |  |  |  |  |  |  |  |  |  |  |
| 2 | 11 | 60 | 0 | 4 | No | 1 | 1 | 85 | Female | High | Yes | No | No | Yes |
| 2 | 11 | 61 | 0 | 2 | No | 2 | 1 | 85 | Female | High | Yes | No | No | Yes |
| 2 | 11 | 62 | 0 | 2 | No | 2 | 1 | 85 | Female | High | Yes | No | No | Yes |

**Null models with random effects terms**

Table S2. Null models with random intercepts at the person and care home levels.

| Null models | Individual level | Care home level | Individual and care home level |
| --- | --- | --- | --- |
| Intercept | 0.001 (0.001,0.001) | 0.001 (0.001,0.001) | 0.001 (0.001,0.001) |
| *Random effects* |  |  |  |
| Individual intercept variance (standard error) | 1.90 (0.08) | - | 0 (0) |
| Care home intercept variance (standard error) | - | 2.67 (0.18) | 2.67 (0.18) |
